# Meta-analysis of the Effect of Orlistat on The Body Mass Index in Obese Subjects

**DOI:** 10.1101/2022.11.28.22282805

**Authors:** Gyeonghwa Jeong, Dongwoo Han, Mahmoud Ahmed, Deok Ryong Kim

**Author notes:** These authors contributed equally (Gyeonghwa Jeong, Dongwoo Han).

## Abstract

Obesity is a widespread health issue worldwide. Therefore, evaluating existing pharmaceuticals and developing new effective strategies to mitigate the problem is essential. Although literature reviews of a broad range of interventions for managing obesity exist, a recent evaluation of the efficacy of orlistat is lacking. This meta-analysis aims to quantify the efficacy of orlistat on body mass index (BMI) and the impact of age, dose, duration, and comorbidities. A literature search of orlistat on PubMed was carried out, and 177 placebo-controlled randomized clinical trials published in the last twenty years were identified. Sixteen studies that met the inclusion criteria were selected for further analysis. Two investigators independently extracted data from the clinical reports using a predefined protocol. We conducted a meta-analysis using random and mixed effects models with different moderators. We found that, on aggregate, the orlistat group reduced their BMI by 0.72 kg/m^2^ (*P* < 0.05) compared to the placebo group. In addition, a longer duration of intervention led to a greater decrease in BMI. Moreover, patients with comorbidities experienced a smaller change in BMI. In conclusion, the evidence suggests that orlistat moderately reduces BMI in obese subjects. The effect of lifestyle modifications, side effects, and drug interactions should be assessed in future studies.

## Introduction

Obesity is a widespread health problem afflicting over 600 million cases worldwide (Ng *et al*., 2014). Therefore, it is of utmost importance that we evaluate existing strategies and develop new ones for managing obesity and its associated comorbidities. Body mass index (BMI) is calculated as weight in kilograms divided by height in square meters and serves as a rule of thumb to classify people’s weight as a function of their height (Yanovski and Yanovski, 2014; Khera *et al*., 2016). US food and drug administration (FDA) approved orlistat for long-term use in obese (BMI ≥ 30) individuals with at least one weight-associated comorbidity (e.g., type-II diabetes, hypertension, hyperlipidemia). The drug functions as a lipase inhibitor, reducing fat uptake, and is currently available as an anti-obesity drug in many countries (Graff *et al*., 2016).

Existing literature reviews have covered various interventions for managing obesity (Singh and Singh, 2020). The effectiveness of Orlistat was also previously reviewed, and it was found to produce superior weight loss compared to a placebo. However, studies evaluating the effects of orlistat are outdated (O’Meara *et al*., 2004). Recent research dealt with a broad range of weight-loss medications but did not specifically focus on orlistat (Singh and Singh, 2020). In this report, we conduct a literature survey and a meta-analysis to evaluate the effect of orlistat on reducing BMI compared to placebo in randomized clinical trials. In addition, we examine the effect of dose, duration of the intervention, and age and comorbidities of the subjects. Finally, we evaluate the quality of existing evidence and the variation between the trials.

## Methods

### Search strategy

A literature search of the electronic database PubMed was carried out on August 1st, 2022. The search term “*orlistat*” in titles and abstracts was used. The following filters were applied to the resulting hits “Species: *humans*,” “Article type: *Randomized controlled trial*,” and “Publication date: *2002/8/1 to 2022/8/1*”. One hundred seventy-seven published reports were identified and reviewed for matching the inclusion criteria. Sixteen studies met all criteria and were included in the meta-analysis. Two independent researchers read and reviewed the reports separately and resolved disagreements by consensus.

### Inclusion and exclusion criteria

Randomized clinical trials comparing the effect of orlistat on weight loss and placebo were retained. Studies using other non-pharmaceutical interventions, such as diet and exercise for weight management, were considered if all the subjects in the control group received the same interventions. The primary measure/outcome was the change in body-mass index (BMI). We chose to use BMI rather than body weight because it is the standard measure for diagnosing obesity (Garrow and Webster, 1985). Also, baseline body weight is different for each participant; it is harder to evaluate the change in outcome with the body weight alone. BMI was presented either by change-score measure or final measure. Studies without data on BMI were excluded. Studies using other weight loss drugs were excluded unless they had a subgroup of orlistat and another control matching non-pharmacological interventions.

### Data extraction

Two investigators extracted the required data from the clinical reports using a predefined protocol. Several data points were extracted from each article: the PubMed ID of the published report, the authors, the year study was published, the region where the study was conducted; the outcome measures; the drug dose, duration of treatment, and non-pharmaceutical interventions (diet, exercise, counseling); the number, age group and, comorbidities of participants. We used as outcomes the mean and standard deviation of BMI change from baseline to end of follow-up. For studies that did not directly report the mean and the standard deviations of change in BMI, we imputed the values by calculating estimates based on the Cochrane Handbook for Systematic Reviews of Interventions Version 5.1.0 (https://handbook-5-1.cochrane.org). In some cases, transformations from confidence interval to standard deviation and final measure into change score were also necessary.

### Statistical analysis

We computed the raw mean difference (MD) and sampling variance of the BMI change from baseline between the treatment and control groups (Borenstein *et al*., 2009). The true variance of the measurements was assumed to be the same in the two groups within each study. To estimate the aggregate effect of the treatment, we used a random effects model with a restricted maximum likelihood estimator (RMEL) (Cooper and Hedges, 1993; Viechtbauer, 2005). Individual coefficients and confidence intervals were based on the normal distribution using a *t*-test. We considered other interventions (dose and duration) and study variables (age group and comorbidities) and estimated their effect on the outcome using a mixed-effects model. Moderators’ coefficients were based on chi-square distribution using the omnibus test.

In both types of models, we estimated the amount of (residual) heterogeneity (Cochran, 1954). Before adding moderators, Q-test was applied to test whether the effect sizes were heterogeneous. After adding moderators to the models, Q_E_-test was applied to determine whether the moderator did not account for the variability in the outcomes. Furthermore, we used the residuals and the standard error in the mixed-effects model to estimate the publication bias (Sterne and Egger, 2001). Finally, we performed a sensitivity analysis to identify influential studies via various leave-one-out diagnostics, including Cook’s distance (Cook and Weisberg, 1982). This analysis was applied using the metafor R package (Viechtbauer, 2010). The source code is available under an MIT license at (https://github.com/BCMSLab/orlistat_meta_analysis).

## Results

### Characteristics of included studies

We identified 177 studies on PubMed, of which only sixteen met the inclusion criteria. Most search results did not report clinical trials and therefore were excluded. Of the ones reporting trial results, the main reasons for exclusion were that the trials were non-randomized, did not include a placebo arm, or included other pharmacological interventions. Several studies were also excluded for not reporting the outcome in terms of BMI. Two articles reported data from the same trial, and therefore oldest of the two was included. Two independent investigators performed the selection and resolved disagreements by consensus.

Among the sixteen studies, five studies were conducted in the US, six studies were in Europe, and studies were in Asia. The size of compared groups (control/comparison) ranged from 12/10 to 265/267. The participants of thirteen studies were adults, and three studies were adolescents. Participants received 120 mg t.i.d orlistat in fourteen studies and 60 mg in two studies. The trials lasted from ten days to a year. Patients also had lifestyle modifications, such as a low-calorie diet, regular exercise, and counseling. Some studies had patients with comorbidities, such as non-alcoholic fatty liver disease, non-alcoholic steatohepatitis, metabolic syndrome, binge eating disorder, impaired glucose tolerance, and hypertension. Table 1. lists the clinical characteristics of the sixteen studies.

**Table 1.** Characteristics of the included studies.

| Author (Year) | Region | N (control/<br>treatment) | Age | Dose<br>(mg, t.i.d.*) | Duration<br>(months) | Lifestyle interventions | Comorbidities |
| --- | --- | --- | --- | --- | --- | --- | --- |
| Esmail VAW (2021) | Iraq | 25/25 | Adults | 120 | 3 | None | Non-alcoholic fatty liver disease, Metabolic Syndrome |
| Shirai K (2019) | Japan | 98/99 | Adults | 60 | 6 | Low caloric diet (reduced 200~400 kcal/day) | None |
| Olszanecka-Glinianowicz M (2013) | Poland | 20/20 | Adults | 120 | 2 | Low caloric diet (1200~1400 kcal/day, low fat), Exercise (5 times/week, 30~40 min; walking, cycling, swimming) | None |
| Yu CC (2013) | Hong Kong | 20/21 | Adolescents | 120 | 2.5 | Low caloric diet (reduced 250~500 kcal/day) | None |
| Carlos M Grilo (2013) | Spain | 19/20 | Adults | 120 | 4 | Nutrition education, exercise, counseling | None |
| Carlos M Grilo (2013) | Spain | 20/20 | Adults | 120 | 4 | Nutrition education, exercise, counseling | Binge eating disorder |
| Smith TJ (2012) | USA | 22/35 | Adults | 60 | 6 | Nutrition, exercise, private counseling | None |
| Harrison SA (2009) | USA | 18/23 | Adults | 120 | 9 | Low caloric diet (1400 kcal/day) | Non-alcoholic steatohepatitis |
| Dixon AN (2008) | UK | 15/16 | Adults | 120 | 12 | Low caloric diet (reduced 600kcal, <30% fat) | Impaired glucose tolerance |
| Ahnen DJ (2007) | USA | 12/10 | Adults | 120 | 1.5 | Low caloric diet (1900kcal, fat 73g) | None |
| Tzotzas T (2007) | Greece | 15/16 | Adults | 120 | 0.33 | Low caloric diet (reduced 1000 kcal/day) | Metabolic Syndrome |
| Audikovsky M (2007) | Hungary | 61/78 | Adults | 120 | 6 | Low caloric diet (1200~1500kcal/day, <30% fat, 55% carbohydrates, 25% proteins) | None |
| Maahs D (2006) | USA | 18/16 | Adolescents | 120 | 6 | Low caloric diet, Exercise, counseling | None |
| Kaya A (2004) | Turkey | 19/25 | Adults | 120 | 3 | Low calorie diet (women 1200, men 1500), Exercise (1 hr walk, fast pace every day) | None |
| Ozkan B (2004) | Turkey | 15/15 | Adolescents | 120 | 12 | Low-caloric diet (20% reduction in daily calories), Exercise (at least 30 min of moderate exercise/day) | None |
| Bakris G (2002) | USA | 265/267 | Adults | 120 | 12 | Low-caloric diet (reduced 600 kcal/day, <30% fat) | Hypertension |

\*t.i.d., three times a day

### Orlistat moderately reduces BMI in obese subjects

First, we sought to evaluate the efficacy of orlistat in reducing body weight in obese subjects. Using a random-effects model, we modeled the mean difference (MD) in BMI between the two experimental groups in the sixteen clinical trials (Table 2, *RE*). The model did not include other variables and treated unknown dissimilarities between the studies as random. We found that, on aggregate, the orlistat group reduced their BMI by 0.72 kg/m^2^ (*P* < 0.05, 95% CI [0.45, 1.00]) compared to the placebo group (Figure 1). Ozkan *et al*., (2004) reported a substantial reduction in BMI (4.20 kg/m^2^, 95% CI [2.27, 6.13]). However, most studies reported a small to moderate benefit, with a wide range of responses between the subjects. We also observed considerable variability between the studies (Q = 39.98, *P* < 0.05), which we attempted to explain using other documented variables.

**Table 2.** Regression parameters and testing output.

| Model | Moderators |  | Test | Heterogeneity (residual) |  |  | Test |
| --- | --- | --- | --- | --- | --- | --- | --- |
| | Term | Estimate (SE) | | $\tau^2$ | $I^2$ | $R^2$ | |
| RE | MD | 0.72 ± 0.15 * |  | 0.15 ± 0.15 | 62.5% |  | Q = 39.98 * |
| ME 1 | Intercept | 0.35 ± 0.25 | $Q_M = 2.82$ | 0.07 ± 0.10 | 29.6% | 34.5% | $Q_E = 25.18 *$ |
|  | Dose (120 mg) | 0.48 ± 0.29 |  |  |  |  |  |
| ME 2 | Intercept | 0.15 ± 0.44 | $Q_M = 6.82$ | 0.09 ± 0.14 | 27.2% | 16.0% | $Q_E = 6.82$ |
|  | Duration | 0.13 ± 0.05 * |  |  |  |  |  |
|  | Age (Adults) | 0.31 ± 0.42 |  |  |  |  |  |
|  | comorbidities (With) | -1.02 ± 0.48 * |  |  |  |  |  |

RE, random-effects; ME, mixed-effects; SE, standard error; MD, mean difference; $\tau^2$ , total amount of heterogeneity; $I^2$ , percent of total variability due to heterogeneity; $R^2$ , amount of heterogeneity accounted for; $Q_M$ , omnibus test; Q, Cochran's Q test; $Q_E$ , Cochran's Q test with moderators. \* indicates $p$ -value < 0.05.

**Figure 1.**
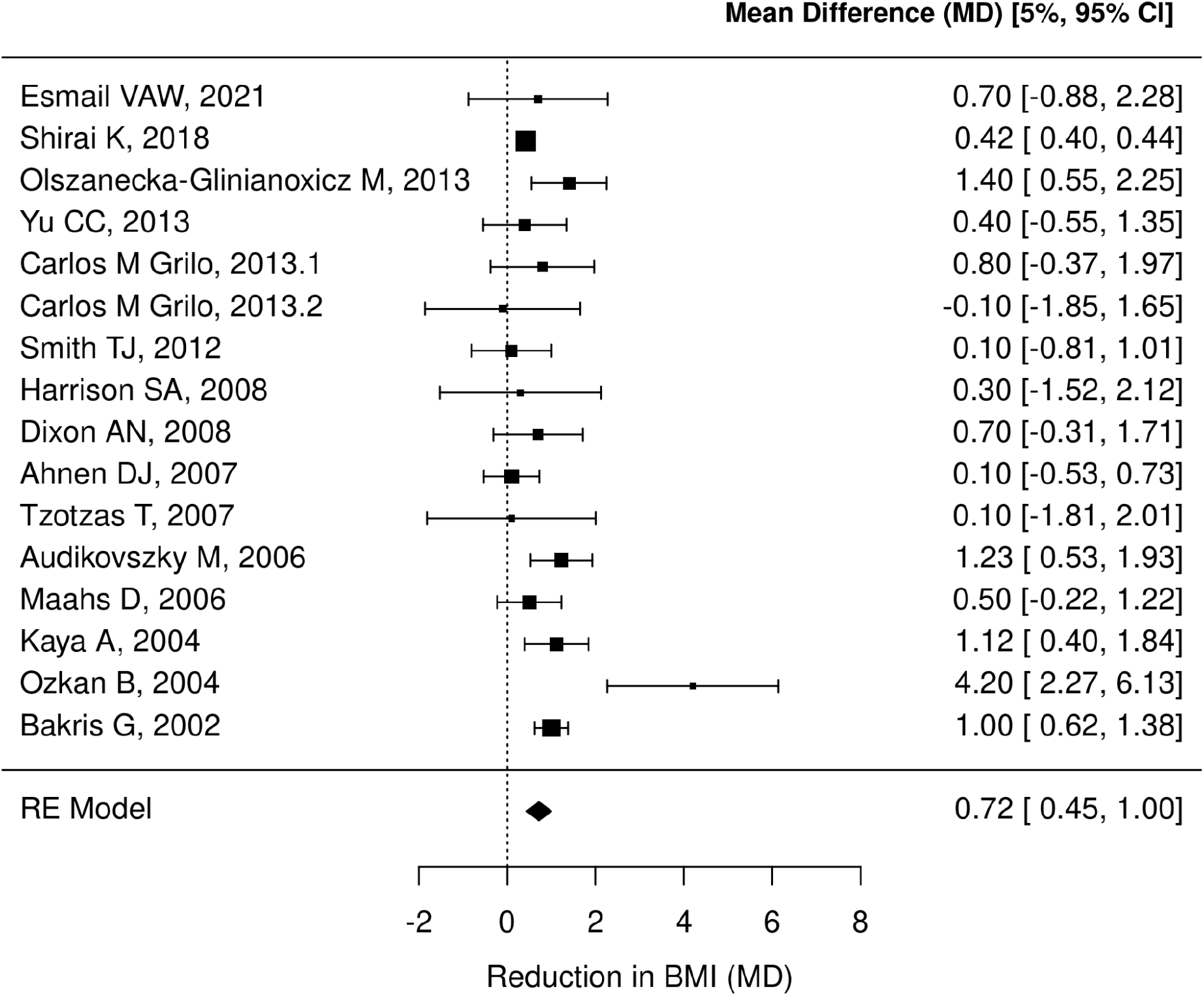
Estimated reduction in BMI in orlistat vs. placebo trials. Included studies (N = 16) are listed on the left by the author(s) and year of publication. Squares represent each study’s mean difference (MD) between treatment and control groups. The squares’ size corresponds to the study’s weight calculated by the inverse of the variance. Lines represent the coverage of the 95% confidence interval (CI). The diamond represents the average mean difference calculated by the random-effects model (Table 2, *RE*). The same values are listed on the right in a numeric form. Positive values mean positive drug effect - decrease on the BMI scale.

### Duration of the intervention, but not the dose, improves the clinical outcomes

We evaluated the effect of the orlistat dose on BMI reduction using a mixed effects model (Table 2, *ME1 &* Figure 2A). We found that when the dose is not considered, using orlistat reduced BMI by 0.35 ± 0.25 kg/m^2^ compared to not using it, but the effect was not statistically significant (*P* > 0.05). Using 120 mg reduced the BMI by 0.48 ± 0.29 kg/m^2^ compared to 60 mg dose, but this was also not statistically significant (*P* > 0.05). The omnibus test of ME1 showed that adding a dose variable was not useful in explaining the final outcome (Q_M_= 2.82, *P* = 0.092) since we could not reject the null hypothesis of the predictor (dose) being unrelated to the outcome. Fourteen studies used a dose of 120 mg, and only two studies used 60 mg. We concluded that the effect of the dose on the BMI reduction could not be estimated, so we excluded studies of the 60 mg dose in the subsequent analysis. We then evaluated the effect of another key study variable, the intervention duration.

**Figure 2.**
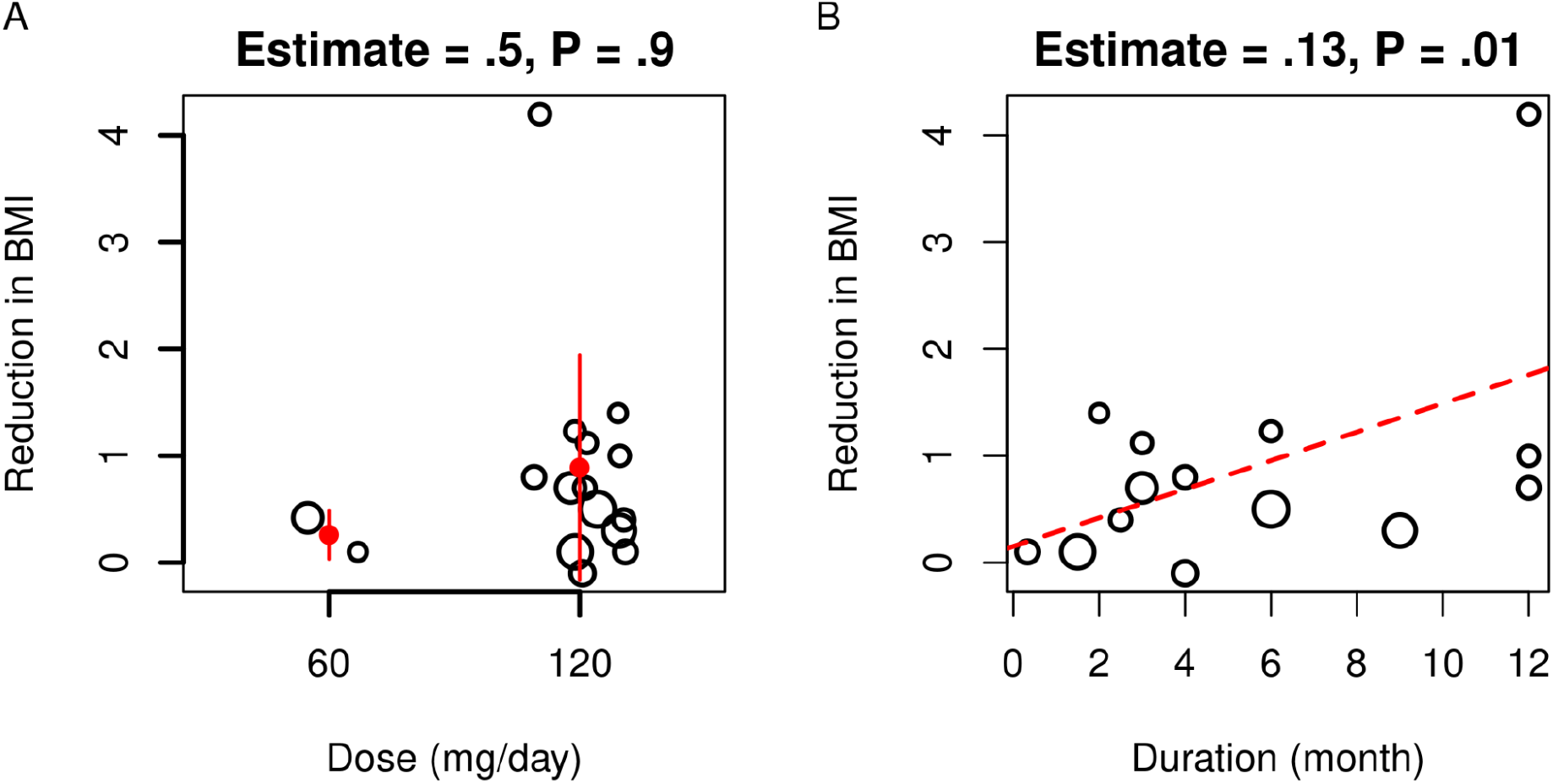
Estimated effects of orlistat dose and duration of treatment on BMI. Each point represents the estimated reduction in BMI units in each study. Point size indicates the standard deviation of the outcome. A) Sixteen studies are stratified into two groups, 60 mg, and 120 mg, and modeled using a mixed effects model (Table 2, *ME1*). Red dots and solid lines are each group’s average and standard deviation. B) Fourteen studies (excluding those with 60 mg dose) with duration (month) on the x-axis and reduction in BMI on the y-axis as modeled using a mixed effects model (Table 2, *ME2*). The dashed red line is the linear trend between the outcome (reduction in BMI) and predictor (duration). Estimate refers to the mean effect of the moderator (dose or duration). P refers to the p-value.

We used another mixed effects model to quantify the effect of the intervention duration, along with the subjects’ clinical characteristics (age group and comorbidities) (Table 2, *ME2*). We found that when duration, age, and comorbidities are held constant, using orlistat reduced BMI by a non-significant 0.15 ± 0.44 kg/m^2^ (*P* > 0.05). However, for each added month, participants reduced their BMI by 0.13 ± 0.05 kg/m^2^ (*P* < 0.05), everything else being the same (Figure 2B). Only some of the predictors were found to be related to the outcome; therefore, the omnibus test was also not significant (Q_M_= 2.82, *P* = 0.077). We then estimated the effects of other study variables on the outcomes.

### Comorbidities impact the response to the intervention

We estimated the differences in outcomes in groups (age and comorbidities) of participants in the mixed effects model mentioned above (Table 2, *ME2*). The difference between adults and adolescents was a non-significant 0.31 ± 0.42 kg/m^2^ (*P* > 0.05) reduction in BMI (Figure 3A). The difference between those who did not have comorbidities and those who did was a 1.02 ± 0.48 kg/m^2^ (*P* < 0.05) reduction in BMI (Figure 3B). Omnibus test of ME2 showed that adding duration, age, and comorbidities variables were not useful in explaining the effect of orlistat on BMI (Q_M_= 6.82, *P* > 0.05). The variability between the studies was a major concern. We next evaluated the effect of these variables on the outcomes and the quality of the evidence.

**Figure 3.**
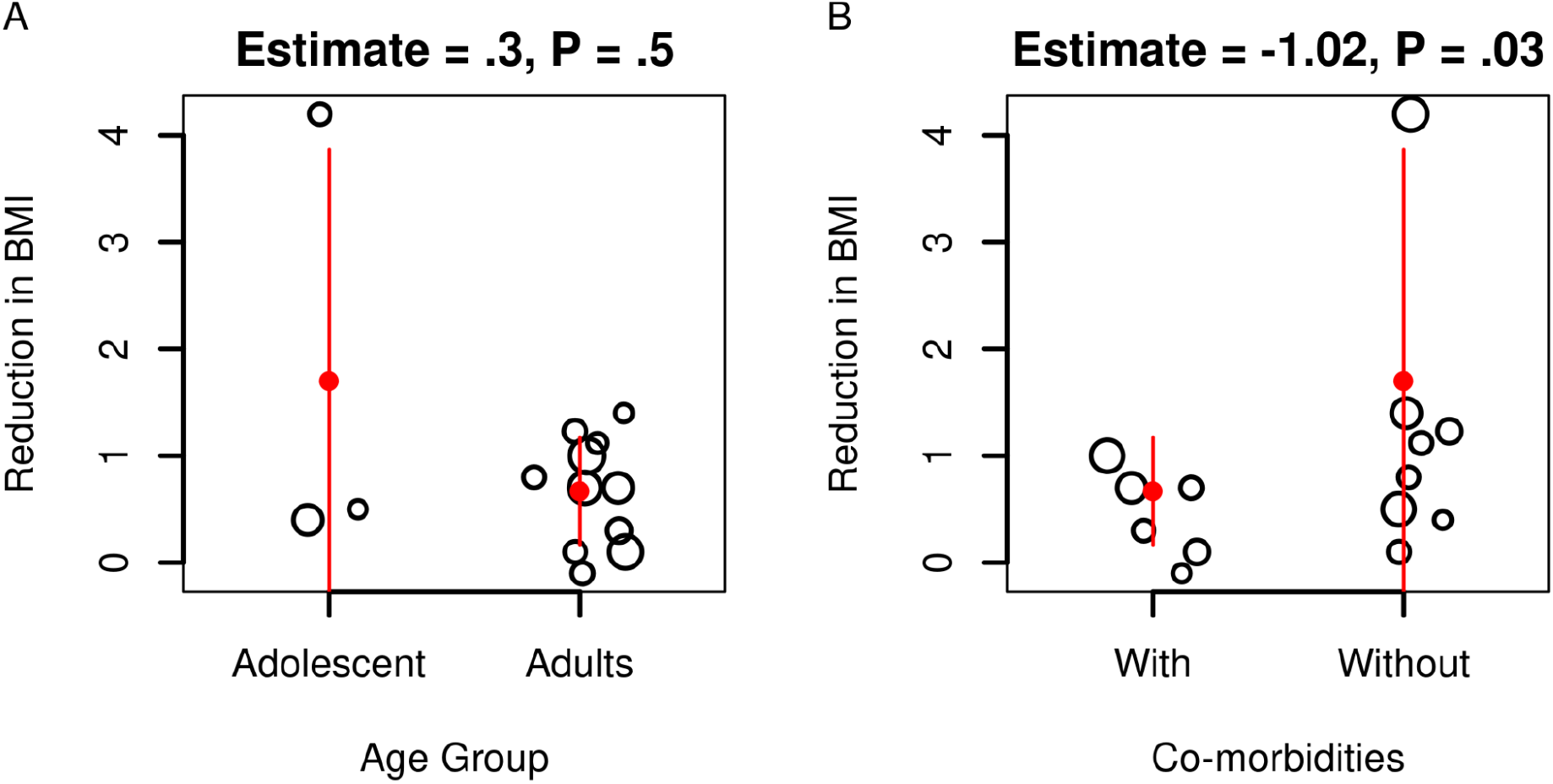
Estimated effects of the age group and comorbidities in the treated subjects. Each point represents the estimated reduction in BMI units in each study (N = 14). Point sizes indicate the standard deviation of the outcome. A) Studies are stratified into two groups; adolescents and adults. B) Studies are stratified into two groups, with or without comorbidities. Red dots and solid lines are each group’s averages and standard deviation. Estimates were calculated using a mixed effects model (Table 2, *ME2*). Estimate refers to the mean effect of the moderator (dose or duration). P refers to the p-value.

### Evaluating heterogeneity, bias, and study influence

We analyzed the heterogeneity among the 16 studies, which we modeled in various ways (Table 2). We considered two measures of variability: **τ**^2^, the total amount of heterogeneity; *I*^2^, the percent of total variability due to heterogeneity; and *R*^2^, the amount of heterogeneity accounted for by including other variables (moderators) in the model. The largest amount of variability was estimated in the random-effects model (RE) (**τ**^2^ = 0.15 ± 0.15), and most of it was due to the heterogeneity (*I*^2^ = 62.5%). The large and significant Cochrane’s Q test (Q = 39.98, *P* < 0.05) indicates larger variation across studies rather than within each study. We considered two fixed-effects modes with different moderators to attempt to explain this variability.

Including the orlistat dose variable in the model (ME1) reduced the total variability (**τ**^2^ = 0.07 ± 0.1) and the amount that is due to residual heterogeneity (*I*^2^ = 29.62%). The drug dose accounted for a significant proportion (*R*^2^ = 34.53%) of the residual heterogeneity between the studies. However, Cochrane’s Q test with age group as a moderator was still large and significant (Q_E_ = 25.18, *P* < 0.05). Meaning more variables need to be considered to explain the across-study variance. The second mixed-effects model (ME2) with duration, age, and comorbidities as moderators had the lowest amount of variability (**τ**^2^ = 0.09) and percent of residual heterogeneity (*I*^2^ = 27.21%). Cochrane’s Q test value was low and insignificant (Q_E_ = 6.82, *P* > 0.05).

Furthermore, we used the residuals and the standard error from the second mixed-effects model (ME2) to estimate the publication bias (Figure 4A). The residual value signifies whether the effects of orlistat are over or underestimated in each study. Five studies overestimated the effect of orlistat, and nine studies underestimated the effect. However, most studies were within the range of expectations. Only Ozkan *et al*., (2004) reported an out-of-range reduction in BMI, with a residual value above 2. So we conclude that there was no significant publication bias.

**Figure 4.**
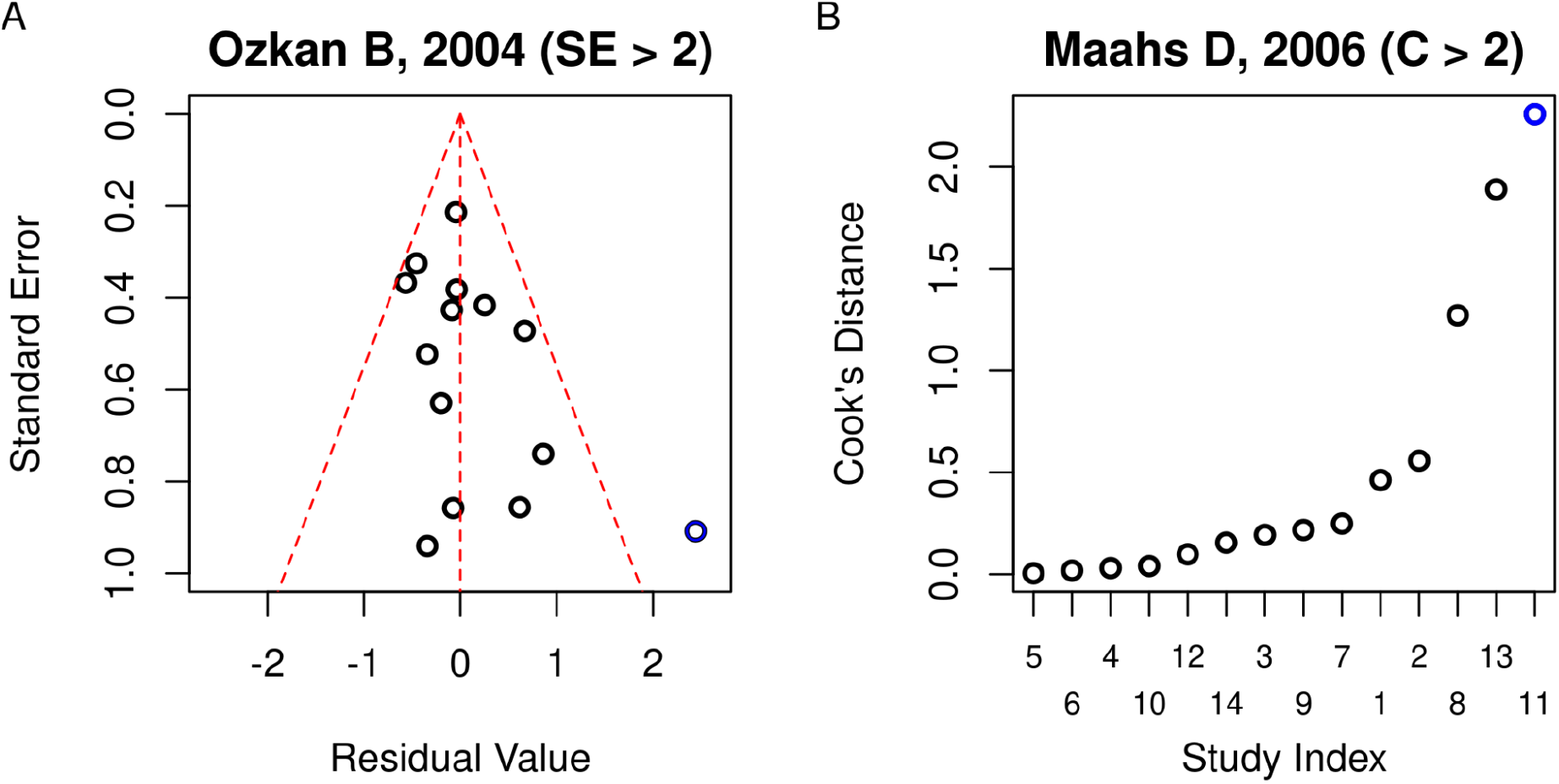
Publication bias and sensitivity analysis of the included studies. Each point represents one study (N = 14). Values are calculated as in the mixed effects model (Table 2, *ME2*). A) A plot of the residual values and standard errors (SE) of the outcomes. B) Studies are indexed as in Table 1 and ordered by Cock’s distance (C). Highlighted studies are outliers.

Finally, we conducted a sensitivity analysis to estimate each study’s influence on the aggregate outcome. We used a leave-one-out strategy to reapply the model and calculate different influence measures. Studies were ordered by Cook’s distance, a measure of how much the regression models differ when the corresponding study is removed (Figure 4B). Most studies had a similar range of significant influence, but three had an outsized influence on the model. Maahs *et al*., (2006) had a Cook’s distance above 2, meaning it significantly influenced the model results.

## Discussion

Orlistat was introduced to reduce weight by inhibiting intestinal fat absorption through inactivating lipase (Graff *et al*., 2016). Our meta-analysis included sixteen randomized clinical trials with 1368 participants, and orlistat can effectively reduce BMI in obese subjects. Our model showed that, on aggregate, the drug decreases BMI by 0.72 kg/m^2^ compared to the controls. A relatively moderate benefit compared to other drugs on the market. A previously published study has shown that sibutramine promoted BMI reduction of -4.2 ± 2.1 kg/m^2^ (*P* < 0.001) compared to orlistat -1.7 ± 0.2 kg/m^2^ (*P* = 0.003) (Anagnostis *et al*., 2012). Another study has demonstrated that liraglutide was more effective than orlistat for weight loss (Astrup *et al*., 2009).

Despite these findings, orlistat can be employed selectively to maximize its benefits. Our analysis showed that with each month of using orlistat, patients had a 0.13 kg/m^2^ decrease in BMI. To our knowledge, This is the first study to quantify the effects of the duration of using orlistat. Patients with comorbidities were less responsive in terms of BMI reduction. However, we could not estimate the effect of different doses due to a lack of studies with diverse dosage regimes.

Several issues with the included clinical reports led to gaps in the analysis and introduced potential inaccuracies. Final outcomes were reported as the final measure or change score, standard deviation, or confidence interval. Therefore, we had to estimate these values in place of the missing data. Next, some studies did not definitively specify whether participants with comorbidities were excluded. Even studies declaring the absence of comorbidities may have comorbidities that were not considered in the clinical characteristics of the cases. Also, most studies did not provide information on ethnicity, only about the region. Finally, many studies assessed the effect of orlistat on weight loss and did not report BMI. As BMI is widely accepted as the standard measure of obesity, we decided to use BMI as our measure (Wharton *et al*., 2020). Because of this choice, we excluded all the studies that only reported weight loss, which resulted in a smaller number of studies for meta-analysis.

Other issues related to the certainty of the evidence arose during this study. Ten studies included zero in their confidence interval, meaning that using orlistat can either increase or decrease BMI. This wide error margin casts doubts on the validity of the evidence, although it may be a result of the natural variation in the participants’ responses. Moreover, the estimated effects of dose and age group on BMI reduction were not significant. Only two studies used a 60 mg dose, which was insufficient to estimate accurately. Although we used different models with moderators to explain the variability and heterogeneity between studies, unexplained residual variation remains.

Resources and the availability of clinical reports constrained our study. First, we could not study the side effects of orlistat, mainly because most studies did not document the side effects of the interventions. Second, the effects of lifestyle changes such as diet, exercise, and counseling could not be estimated. Third, we selected provided the same exercise, diet, and counseling in the placebo and treatment groups. Fourth, most excluded reports did not provide information on the change in BMI from the baseline. BMI is widely accepted as the standard measure of obesity, so we decided to use BMI as our outcome and exclude the studies that only provided weight information. Finally, this study did not evaluate the interactions between orlistat and other drugs. Most studies did not report information on drug interaction, and we could not collect enough data for analysis.

## Conclusions

In conclusion, the present data suggest that orlistat moderately reduces BMI in obese subjects. Moreover, using orlistat for longer periods has a greater effect. Orlistat is less effective in cases with comorbidities. More studies are needed to confirm the effect of the dose of orlistat. Future studies should be designed to investigate the effect of lifestyle modifications, the side effects, and the interactions of orlistat with other drugs.

## Data Availability

All data produced in the present work are contained in the manuscript.

https://github.com/BCMSLab/orlistat_meta_analysis

## Declarations

## Acknowledgments

This study was supported by the National Research Foundation of Korea (NRF) grant funded by the Ministry of Science and ICT (MSIT) of the Korea government [2020R1A2C2011416]. GJ and DH were part of a training program for medical students funded by the National Health Insurance Service in Korea.

## Author contributions

GJ and DH conducted the literature survey and data extraction and contributed to the analysis and writing of the manuscript. MA contributed to the analysis and writing of the manuscript. DRK acquired the funding, supervised the project, and contributed to the writing of the manuscript.

